# Targeting a future generation free from female genital mutilation: a mixed-methods quasi-experimental study of an awareness intervention in central Tanzania

**DOI:** 10.1101/2025.09.26.25336717

**Authors:** Leah Barthalome Kimario, Manji Nyaganya Isack, Beatrice Temba, Agnes Cyril Msoka, Blandina Theophil Mmbaga

**Author notes:** **Corresponding author** (LBK).

## Abstract

Female genital mutilation (FGM) persists in Tanzania, with the Dodoma Region having the country’s second-highest prevalence. This study evaluated a community-based educational intervention to increase FGM awareness and promote abandonment among young adults (15-19 years) in Chamwino District, Dodoma, a key but often neglected group for fostering intergenerational change. We conducted a mixed-methods study with a primary quasi-experimental component: a single-arm, baseline-endline survey with a multi-stage cluster sample of 452 completed follow-up young adults from schools and hospitals. Secondary components included a clinical audit of delivering mothers to determine FGM prevalence and in-depth interviews with ten FGM-positive young mothers. The primary outcome was the change in the proportion of participants with “Adequate Awareness” of FGM health risks; secondary outcomes included changes in attitudes and observed prevalence. Quantitative data were analyzed using paired t-test, McNemar’s test, chi-square, and logistic regression, reporting Adjusted Prevalence Ratios (aPRs), odds ratios, or mean differences with 95% Confidence Intervals (CIs), as appropriate. Qualitative data were thematically analyzed. The observed FGM prevalence was 16.6%. Adequate awareness increased from 1.3% to 14.6% (aPR = 5.45, 95% CI: 4.62–6.29, p<.001). Recognition of FGM as harmful rose from 91.2% to 96.7%, and the desire for its abandonment increased from 81.0% to 95.8% (both p<.001). Qualitative analysis yielded three themes: 1) The hidden system, 2) Blood and lies, and 3) Intergenerational revolt. A theory-informed, community-engaged intervention significantly improved knowledge and attitudes among more than 1,700 community members. However, qualitative findings revealed a persistent “hidden system” and a “critical power gap” among youth, indicating awareness alone is insufficient. Sustainable abandonment requires integrating awareness campaigns with strategies that address structural power dynamics.

## Introduction

FGM affects over 230 million women and girls globally, mainly in Africa [1], with 8% of reproductive women in Tanzania affected [2]. FGM-related complications are severe and range from short-term to long-term, including urogynecologic, psychological, and obstetric complications [3], requiring ongoing efforts undertaken by this study to protect the next generation of women. The Dodoma Region has the country’s second-highest FGM prevalence (47%), and practices are shifting to early childhood to evade laws [2,4]. FGM endures due to deep-rooted socio-cultural norms and gender inequalities, despite laws and awareness efforts [5]. A critical oversight is the limited focus on adolescents and young adults, who are influential, less bound by tradition, and more receptive to change [6,7]. Empowering them supports Sustainable Development Goal target 5.3 to eliminate harmful practices [8].

This gap is especially evident in high-prevalence, rural districts like Chamwino in Dodoma, a Gogo tribe heartland with high FGM rates [9,10]. Despite this, little is known about young adults’ awareness, attitudes, and experiences regarding FGM in Chamwino, and no youth-focused programs have been documented. This study aimed to evaluate a theory-informed, community-based educational intervention designed to increase awareness of FGM risks and promote abandonment among young adults. We hypothesized that participants in the multi-component educational intervention would demonstrate a significant increase in adequate awareness of FGM health risks compared to their baseline awareness. Using mixed methods, we also explored the FGM prevalence and experiences to contextualize the impact and identify persistent socio-structural barriers to abandonment.

## Materials and methods

### Study design and registration

This was a community-based, mixed-methods study from April 2023 to March 2024. The primary component was a single-arm, baseline-endline quasi-experimental design (non-randomized) to evaluate a multi-component educational intervention. The other components were: a prospective clinical audit of FGM prevalence to provide contextual depth; qualitative interviews to explain the quantitative findings; and a process evaluation of implementation feasibility and reach. The study protocol was finalized before participant enrollment and was retrospectively registered with the Open Science Framework (OSF) [https://doi.org/10.17605/OSF.IO/XCQRT] to ensure transparency. Reporting of quantitative findings followed the Transparent Reporting of Evaluations with Nonrandomized Designs (TREND Statement Checklist) [11] and qualitative findings followed the Consolidated Criteria for Reporting Qualitative Research (COREQ) [12]. The study proceeded in four phases: (1) Preparation and baseline (April-June 2023): obtaining approvals, validating research/intervention instruments, and starting longitudinal data collection with a baseline survey and prevalence data; (2) Intervention initiation (June-August 2023): training community champions, forming an evaluation team, and starting qualitative interviews; (3) Implementation and monitoring: deploying intervention activities across schools, hospitals, and communities while continuing prevalence and qualitative data collection; and (4) Endline and evaluation (March 2024): endline survey, final analysis, and community feedback.

### Theoretical framework

The intervention integrated behavioral models to address the multifaceted drivers of FGM. The Health Belief Model shaped perceptions of health risks and perceived benefits of abandonment. Social Cognitive Theory employed peer champions and testimonials to build self-efficacy and model new norms. The Theory of Planned Behaviour targeted attitudes, subjective norms (social pressure), and perceived control. The Socio-Ecological Model structured the intervention across levels: individual (knowledge), interpersonal (family dialogues), community (mobilization), and societal (laws and gender norms) [13–16].

### Study setting and participants

This project was conducted in Chamwino District, a high-prevalence area of FGM in the Dodoma Region of Central Tanzania [2,10]. Besides the Gogo, FGM in Chamwino is practiced across multiple tribes, including both indigenous and migrant groups such as Sandawe, Burunge, Rangi, Maasai, and many others. FGM remains discreet, with targeted interventions lacking in key groups like young adults. In 2022, Chamwino’s population accounted for approximately 16% of Dodoma’s population (486,176 of 3,085,625), including 49,807 young adults [17]. Six of 36 district wards were purposively selected to ensure urban/rural diversity, covering known variations in FGM prevalence and practical considerations related to budget, timeline, and geographical accessibility, given the district’s ward dispersions. We aimed to include one secondary school (with at least 71 students aged 15-19) and one public health facility (with at least seven deliveries among young adults in the past three consecutive months) to facilitate rapid recruitment. Low delivery rates at Itiso health facilities and during the holidays in the baseline survey at Mlowa and Mpwayungu Wards resulted in respective schools lacking samples. Missed samples were evenly distributed across similar strata (e.g., school-for-schools, hospital-for-hospital), and analysis was adjusted accordingly. The final sample (5 hospitals, four secondary schools), with their corresponding baseline-endline survey participants, is detailed in Table 1.

**Table 1.** Study wards, participating institutions, and corresponding baseline-endline participants.

| Ward | Type | Representatives |  |  |  |  |  |
| --- | --- | --- | --- | --- | --- | --- | --- |
|  |  | Health Facility | Secondary school | Participants/Ward |  |  |  |
|  |  |  |  | Baseline |  | Endline |  |
|  |  |  |  | M | F | M | F |
| **Mpwayungu** | Remote | Mpwayungu Health Centre | — | — | 8 | — | 8 |
| **Haneti** | Remote | Haneti Health Centre | Haneti | 59 | 57 | 57 | 53 |
| **Chamwino** | Semi-urban | Chamwino Health Centre | Chamwino | 59 | 57 | 59 | 56 |
| **Itiso** | Remote | — | Itiso | 58 | 48 | 54 | 46 |
| **Mlowa** | Semi-urban | Chamwino District Hospital | — | — | 8 | — | 8 |
| **Dabalo** | Remote | Dabalo Health Centre | Dabalo | 58 | 56 | 58 | 53 |
| **Total** |  | **5** | **4** | **234** | **234** | **228** | **224** |

### Participants’ sampling, recruitment, and eligibility criteria

Participant recruitment spanned 10 months, from the baseline survey (May 26–June 9, 2023) to the endline (March 1–15, 2024). FGM prevalence data were collected consecutively for 9 months (June 1, 2023–February 29, 2024), and qualitative interviews over 6 months (July 30, 2023–January 16, 2024)—sampling methods varied by component.

Baseline-endline participants were young adults aged 15–19 enrolled in four schools or receiving maternity services at five hospitals. To minimize loss to follow-up, individuals planning to leave the district and those in their final year of schooling were excluded, alongside individuals unable to consent. For young mothers, baseline data were collected during the postnatal hospital stay. Given the practical difficulty of tracking participants after discharge, follow-up strategies included linkage to scheduled postnatal visits and optional, participant-defined contact methods to facilitate endline data collection.

Multi-stage cluster sampling with probability proportional to size was used, assuming a 10:1 school-to-hospital eligible population ratio. The feasibility-based target (468 calculated sample) was 426 from schools and 42 from hospitals, balanced by gender and ward (78; 71 from schools and seven from hospitals). To meet the school target amid an anticipated ∼30% non-response rate due to a sensitive topic, consent/assent forms were given to 610 students in randomly selected classes across four schools; 510 (83.6%) returned. After verification, 42 students were excluded (age outside range: n=18; residency <1 year: n=16; other: n=8), 426 were enrolled, matching the school target precisely. In hospitals, census sampling was used. No refusals were recorded among the 42 young mothers delivering at baseline who were approached. Total baseline enrollment was 468, meeting the target, with 452 (96.6%) retained for paired analysis.

Qualitative purposive sampling selected 10 FGM-positive young mothers (15–19) who recently delivered at study hospitals, knew of ongoing FGM, and were willing to share experiences. This sample size, within a recommended range (5-25) for phenomenological studies [18] allowed in-depth exploration until thematic saturation at the phenomenon level [19]. Four declined due to fear. Those unable to consent, in pain, or unable to endure post-birth interviews were excluded.

The FGM prevalence audit included all delivering mothers at five hospitals. Over 9 months, 3,842 mothers delivered; 72 (1.9%) were excluded (missing consent: n=48; refusal: n=24), leaving 3,770 (98.1%). General inclusion and exclusion criteria for the baseline-endline survey and interview: participants had to have lived in the district for at least 1 year, ensuring sufficient exposure. Those unable to consent, cognitively impaired, or with less than a year of residence were excluded. The participant flow is detailed in Fig 1.

**Figure 1.**
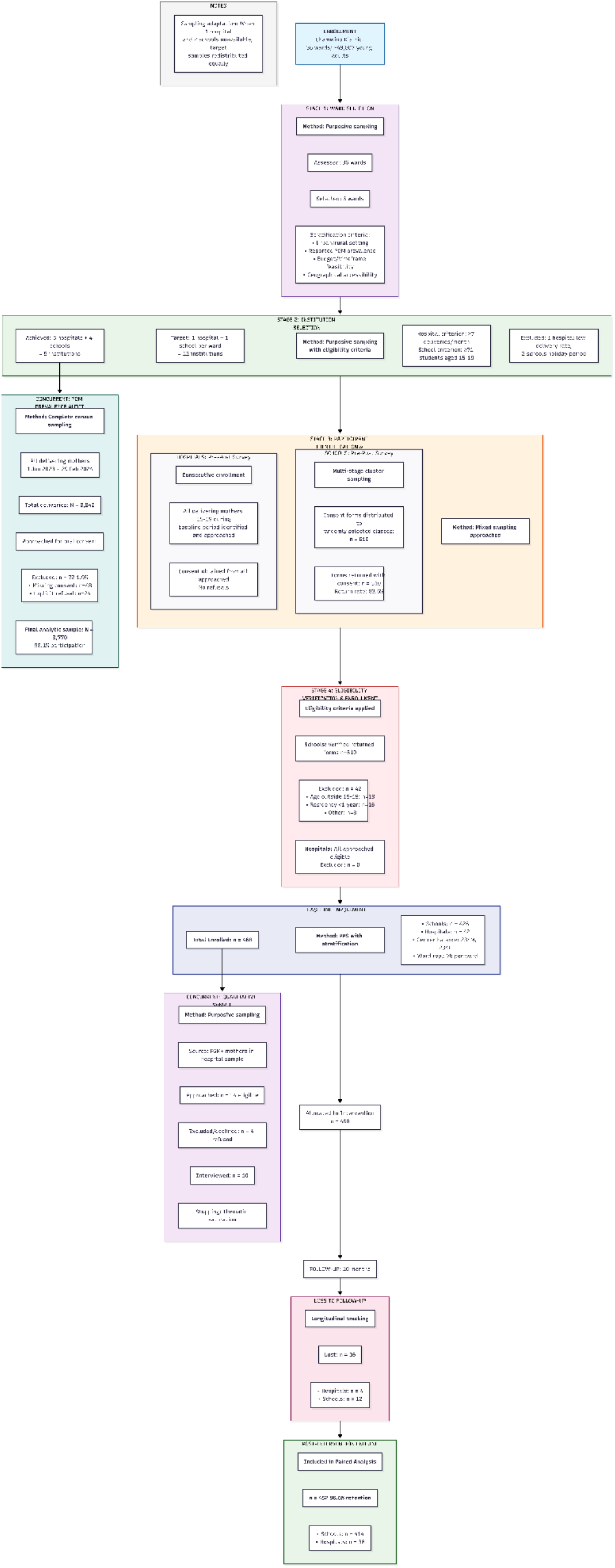
**Participant flow and sampling diagram for the mixed-methods study**

### Interventions

The intervention was a multi-component, community-based educational campaign, following the literature-based training manual (S4 File) [2,20–23], informed by the four integrated behavioral theories [13–16]. These theories served as independent variables in the quasi-experimental design, allowing empirical assessment of their causal impact on FGM awareness and attitudes among a previously unresearched, intervention-naïve population.

Fifty champions, 15 adults from (health workers, teachers, local/religious leaders, elders, traditional birth attendants, community health workers), and 35 students were trained. An 11-member evaluation team, including the two lead authors, representatives from the champion groups, and a police officer, had three key roles: (1) validated data collection instruments, training manual, and intervention activities for cultural and contextual fit, (2) monitored the intervention’s impact, and (3) set sustainable goals. Implementation of activities was ward-based, guided by baseline needs, and adapted to the district’s dispersed geography. The core educational content from the validated manual was consistently implemented.

Educational sessions were delivered across multiple settings. In schools, 737 students participated in initial gender-stratified sessions. Student and teacher champions collaborated to establish health clubs and debate sessions. Health clubs, with 35 participants (totaling 105 attendances), met three times at one school, and four debate sessions were held in two schools without gender stratification to promote shared perspectives. Attendance at the open-access debate sessions was not formally recorded because they were voluntary forums for open dialogue, prioritizing discussion over participant counts. In hospitals, 13 training sessions were delivered in labor wards and antenatal clinics to 246 participants. Nurse-midwives received additional training on the World Health Organization (WHO) FGM typology [20,24] using visual aids to improve clinical documentation. Approximately 80% of nurse-midwives were trained to maximize data capture across hospitals.

Community mobilization involved household visits and large gatherings. Guided by feasibility estimates, a team of 15, including 10 champions (7 students, 3 teachers) and the two lead authors, conducted 46 household visits over a month in one ward and two large gatherings in two wards, thereby facilitating broader open dialogue. A local government representative accompanied all activities. All household members were invited, but the census was only for those 12 or older who could comprehend the educational message [25]. No households refused consent.

### Outcomes and measurement

The primary outcome was the change in the proportion of participants with ‘Adequate Awareness’ of FGM health risks, defined as a score >76% on a 19-item knowledge scale, with scores categorized as inadequate (<50%), moderate (50–75%), and adequate (>76%) [26]. Mean awareness score was examined as a continuous measure to provide further detail on awareness gains. Secondary outcomes included (1) changes in attitudes: the proportions of participants who believed FGM is harmful, acknowledged its continuation in the community, and desired its abandonment, (2) the observed prevalence of FGM among delivering mothers, and (3) process measures of intervention reach and fidelity.

To enhance data quality, the three Swahili-language instruments were developed from the literature [2,5,7,20,22,23]. All instruments were pretested and refined in a non-participating ward among the eligible samples. The 19-item awareness scale demonstrated acceptable internal consistency in our baseline sample (Cronbach’s alpha = 0.70) and was then pretested with 47 secondary school students (10% of 468). The interview guide was tested through two hospital interviews. The FGM audit checklist was briefly piloted with labor ward staff (10 cases) to ensure accurate categorization and workflow integration.

### Qualitative component

A concurrent qualitative phenomenological design was used to explore the lived experiences, perceptions, and contextual barriers among FGM-positive young mothers. The purpose was to understand the physical, psychological, and social impacts from the perspective of those who have undergone the practice, and explain the quantitative findings in this previously unexplored community.

### Data collection procedures

Data collection procedures were tailored to each component. For the survey, administration (self- or interviewer-based) was adapted to literacy levels. School-based surveys were administered in open areas, with participants spaced 1 meter apart, to reduce social desirability bias and ensure privacy; completeness was verified on-site. For FGM prevalence, trained nurse-midwives used a standardized checklist to collect primary data from all delivering mothers during care prospectively. Data quality was ensured through cross-verification against birth registries, specialized training in the WHO FGM typology, and supervision. For qualitative, the two lead authors (LBK, MNI), motivated by FGM abandonment, bearing clinical and public health experience, conducted separate five private, face-to-face, audio-recorded in-depth interviews each in Swahili with FGM-positive young mothers in hospital labor wards, to reduce bias. Interviews lasted 30–60 minutes, followed member-checking procedures, and were supplemented with field notes. Interviews were once conducted without repeats or a third person present.

### Data analysis

Quantitative analysis used SPSS v25 to evaluate intervention effects, providing detailed data on participant demographics and outcome distributions to inform stakeholders and guide future strategies. The primary analysis followed a complete-case approach (n=452). The change in the mean awareness score (primary continuous outcome) was assessed with a paired t-test after confirming normality (Shapiro-Wilk test: W = .995, p = .130).

Intervention effect sizes on changes in awareness levels (primary categorical outcome) were estimated using aPRs with 95% CIs via Modified Poisson Regression with robust error variances, providing a direct, interpretable estimate of effect for common categorical outcomes, rather than risk or odds ratios [27]. Endline prediction of participant demographics to the primary outcome was assessed via multinomial logistic regression.

Changes in secondary binary outcomes were analyzed using McNemar’s test, and associations with participant demographics were examined using chi-square tests and binary logistic regression, as appropriate. Descriptive statistics summarized FGM prevalence. All inferential statistics employed a 5% significance level and reported both p-values and effect measures (mean differences, aPRs, or odds ratios, as appropriate) with 95% CIs, per contemporary reporting standards [28].

For the qualitative data, audio recordings were transcribed verbatim, translated into English, and analyzed manually following Braun and Clarke’s inductive thematic analysis framework [29]. To ensure analytical rigor and mitigate bias, LBK and MNI first independently familiarized and coded the same five transcripts. They then met in a series of discussions to compare, reconcile, and refine their codebooks into a consensus-based preliminary thematic framework. This framework was applied to the remaining transcripts, refined collaboratively, then reviewed and validated by the entire research team.

Quantitative and qualitative findings were integrated via narrative synthesis in the Discussion.

## Results

### Intervention reach, fidelity, and process evaluation

The campaign directly engaged over 1,700 individuals, including more than 900 students, 246 in hospital settings, 104 in households, and over 500 attendees at the two community gatherings. Process evaluation indicated strong implementation fidelity and high participant engagement, with trained champions leading activities. Student champions served as change agents, serving as main speakers during household visits and community gatherings to promote local ownership and message credibility, and were briefed in advance on the content and language. Health workers and traditional birth attendants shared testimonies in health clubs, with a police officer providing legal and reporting mechanisms. Anonymous feedback showed high project ownership with a mean proportion (84.7%) across 77.9% (household visits), 83.6% (community gatherings), and 92.5% (endline survey).

Health clubs were sustained post-study and evolved into broader health education platforms. Furthermore, a private WhatsApp group connected dispersed adult champions for remote supervision and peer support. Following the study, this developed into a sustained, community-led network for ongoing anti-FGM initiatives. Student champions, excluded due to school phone policies, were integrated through in-person meetings and final planning sessions.

### Endline participants’ demographics

Among the 452 who completed follow-up, the median age was 17 years, 50.4% were male, and 93.1% had secondary education. Chamwino Ward contributed the most significant number of participants (25.4%). Most (72.8%) were from remote wards, and 91.6% were recruited from schools. Of the 33 participant tribes, the majority were Gogo (57.5%). The median community residence was 16 years (Tables 1 and 2). Of the 468 young adults at baseline, none had prior anti-FGM awareness program participation (S1 Table), confirming a critical gap in targeted interventions, which the present study aimed to address.

**Table 2.**
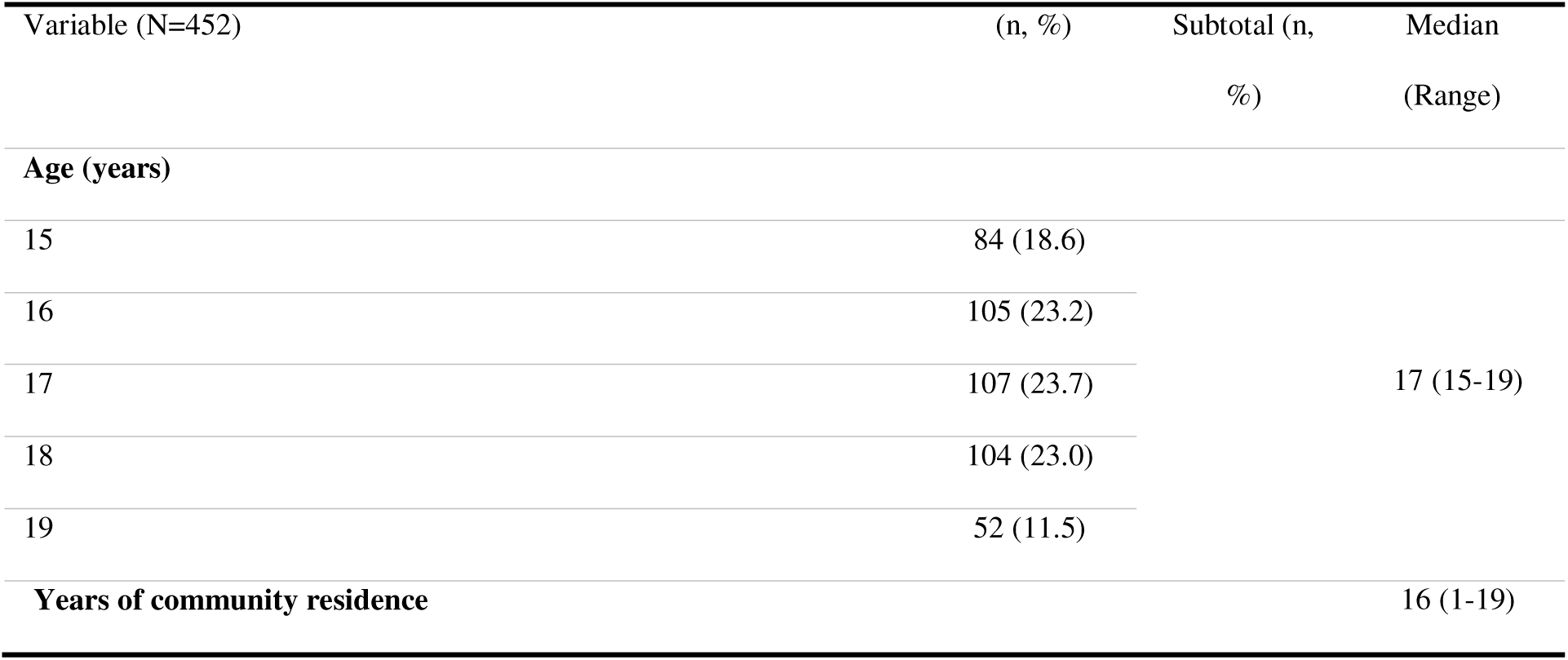
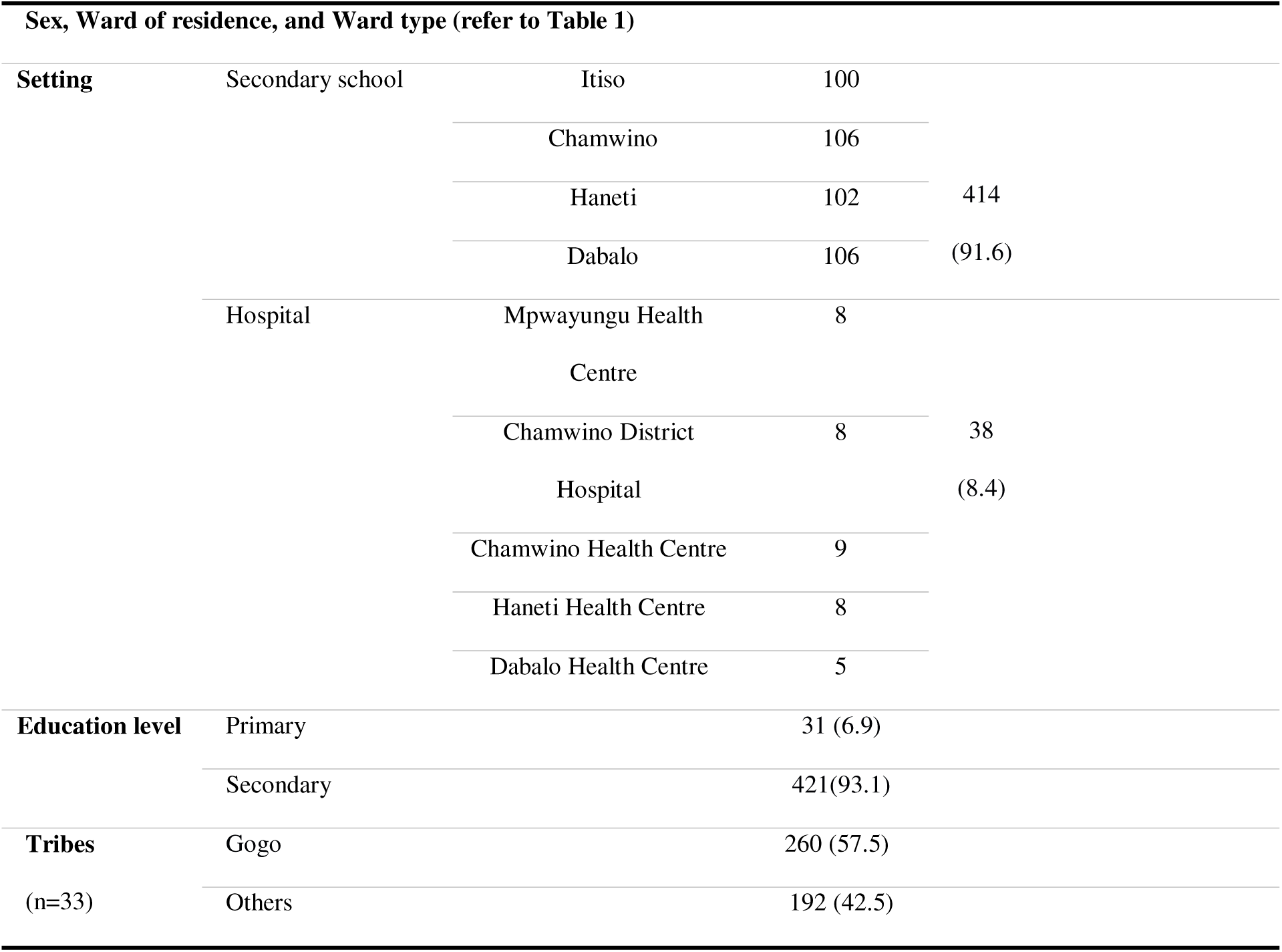
Socio-demographic characteristics of young adult participants (N=452) in the endline survey, Chamwino District, Tanzania.

### Attrition analysis

Analysis of baseline characteristics between participants retained for analysis (n=452) and those lost to follow-up (n=16) showed no statistically significant differences (all p > 0.05) in sex, age, ward of residence, type of ward, tribe, education level, self-reported FGM status among females, or, most importantly, mean awareness score. Attrition differed by setting (p < 0.001), with more loss from schools (12/16) than hospitals, consistent with the greater logistical difficulty of tracking school-based participants over time. Overall, attrition was minimal (3.4%) and unlikely to have introduced systematic bias in the primary findings [30].

### Intervention impact on FGM awareness and knowledge

FGM adequate awareness increased from 1.3% to 14.6% (aPR = 5.45, 95% CI: 4.62, 6.29; p < .001) (Fig 2). The mean awareness score increased by 15.04 points (95% CI: 12.90, 17.18), from 44.08 (SD = 15.85) to 59.12 (SD = 16.85); t(451) = 13.81, p < .001. Among the participant characteristics, only ‘type of ward’ was statistically significant in the adjusted model (p < 0.05). Participants from semi-urban areas had higher odds of achieving moderate awareness (aOR = 3.30, 95% CI: 1.02, 10.66; p = .046) and adequate awareness (aOR = 9.26, 95% CI: 2.30, 37.20; p = .002) than those from remote wards.

**Figure 2.**
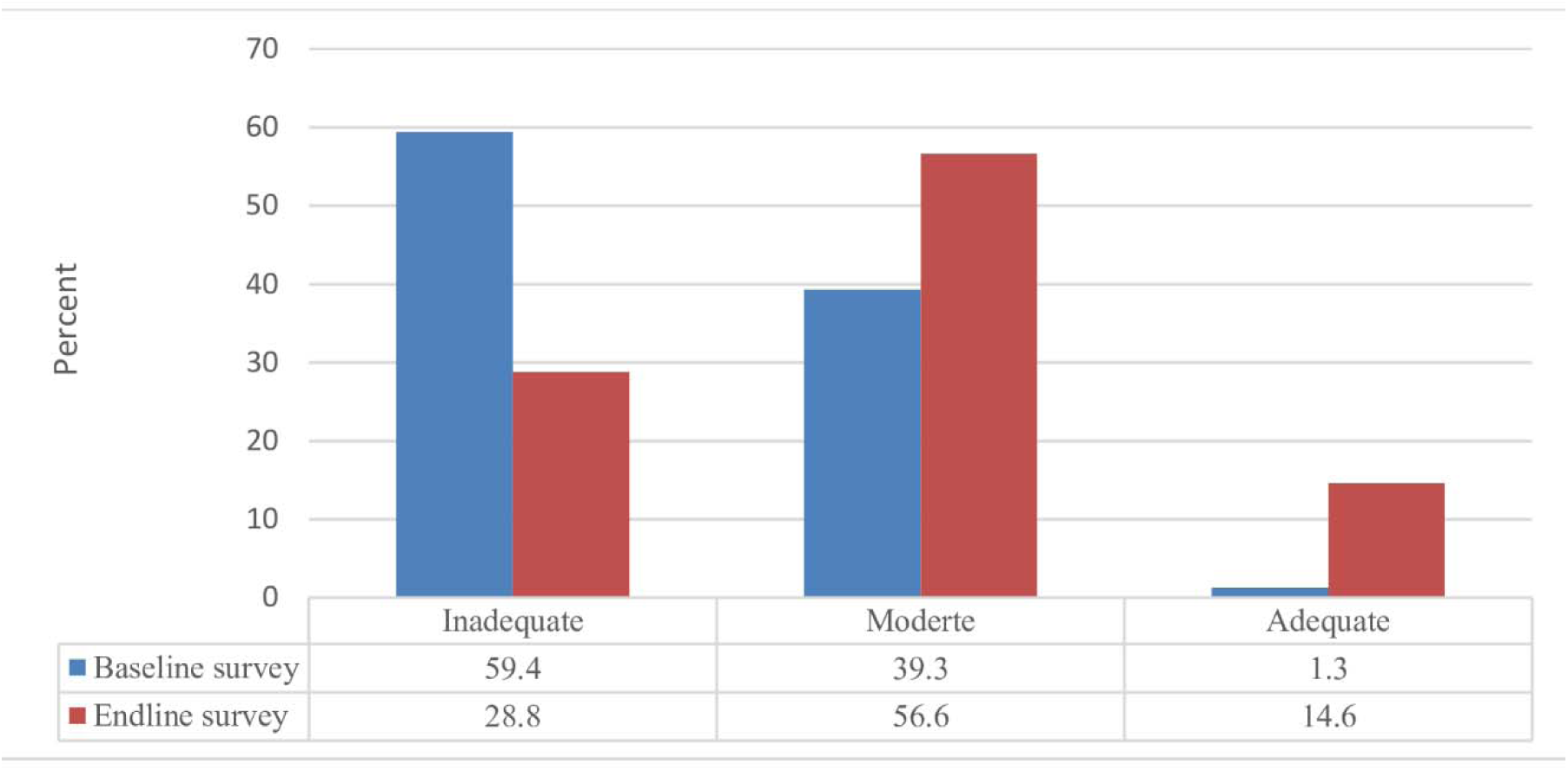
**Distribution of FGM awareness levels among young adults before and after a community-based educational intervention, Chamwino District, Tanzania**

### Changes in attitudes, perceptions, and intention to abandon

Recognition of FGM as harmful increased from 91.2% to 96.7% (McNemar’s χ² = 10.47, p = .001), a net increase of 25 individuals (40 gained awareness, 15 regressed). Participant characteristics were not significant predictors of this change (all p > 0.05). At endline, the most commonly identified health consequence was excessive bleeding (91.5%). Furthermore, recognition of FGM as harmful was weakly associated with familiarity with its ongoing practice in the community (χ²(1) = 5.84, p = .016, φ = .114), but not associated with the desire for its abandonment.

At endline, knowledge improved sharply: recognition that FGM lacks medical justification rose from 52.6% to 92.7%, and awareness of its criminal status increased from 87.3% to 96.7%. Acknowledgment of FGM as a human rights violation rose from 85.0% to 97.6%; among those recognizing it as a violation, the most cited associated abuses were child/early/forced marriage (61.7%) and intimate partner violence (58.4%). Understanding of the ‘World’s FGM Day’ increased from 42.3% to 95.8%. School education was the primary reported source of FGM awareness (93.1%), followed by mass media (29.2%) and community discussions (16.2%) (multiple responses allowed).

Acknowledgment that FGM is still practiced in the community increased from 39.7% to 50.9% (McNemar’s χ² = 9.65, p = .002), a net increase of 51 individuals (155 changed from ‘No’ to ‘Yes’, 104 regressed). Male participants had 2.35 times the odds of acknowledging its continuation compared with females (95% CI: 1.44, 3.83; p = .001), whereas longer community residency was associated with 6% lower odds (aOR = 0.94 per additional year, 95% CI: 0.89, 0.99; p = .012). Among those aware, 60.9% could not specify reasons for its persistence, although preserving norms and honoring ancestors were the most frequently cited reasons (17.4%) (Table 3).

**Table 3.** Perceived reasons for the continuation of FGM among participants aware of its.

| Reason | n=230 (%) |
| --- | --- |
| Enhancing women's value during marriage | 5 (2.2) |
| Preventing stigma/earning respect | 4 (1.7) |
| Controlling women's sexuality | 13 (5.7) |
| Preserving norms and honoring ancestors | 40 (17.4) |
| Children's inability to question elders | 2 (0.9) |
| Rite to womanhood | 6 (2.6) |
| Treating vagina and urinary tract infections ("lawalawa") | 3 (1.3) |
| Do not know/ non-response | 140 (60.9) |
| Insufficient education on consequences | 14 (6.1) |
| Lack of community-tailored anti-FGM programs | 3 (1.3) |

The desire to abandon FGM increased from 81.0% to 95.8% (McNemar’s χ² = 40.63, p < .001), a net increase of 66 individuals (85 changed from ‘No’ to ‘Yes’, 19 regressed). This positive attitude was nearly universal at endline, with no significant differences across participant characteristics. Table 4 stratifies the key endline findings by participant sex.

### FGM prevalence and typology

A nine-month prospective audit across five hospitals included 3,770 delivering mothers aged 13-41. The overall observed FGM prevalence was 16.6% (624/3770), higher among young mothers aged 15-19 (31.0%, 158/510). Most FGM cases were Type II (97.4%) and among the Gogo tribe (73.6%). The remote Haneti Ward had the highest FGM cases, at 51.5% (270/524), equivalent to 43.3% (270/624) of all cases in Chamwino District. Only 7.3% (17/234) of women self-reported FGM-positive status in the baseline survey.

**Table 4.** Key endline findings, stratified by participant sex (N = 452)

| Variable |  | Male<br>(n=228) | Female<br>(n=224) | (n, %) |
| --- | --- | --- | --- | --- |
| FGM awareness<br>level | Adequate | 35 | 31 | 66 (14.6) |
|  | Moderate | 132 | 124 | 256 (56.6) |
|  | Inadequate | 61 | 69 | 130 (28.8) |
| FGM is harmful |  | 223 | 214 | 437 (96.7) |
| Harmful effects of FGM (Multiple responses allowed) | Excessive bleeding | 207 | 193 | 400 (88.5) |
|  | Severe pain | 184 | 173 | 357 (79.0) |
|  | Infections | 166 | 145 | 311 (68.8) |
|  | Urinary problems | 125 | 137 | 262 (58.0) |
|  | Psychological problems | 118 | 113 | 231 (51.1) |
|  | Sexual problems | 130 | 122 | 252 (55.8) |
|  | Difficult childbirth | 140 | 138 | 278 (61.5) |
| Sources of FGM awareness (Multiple responses allowed) | School | 213 | 208 | 421 (93.1) |
|  | Mass media | 71 | 61 | 132 (29.2) |
|  | Community discussions | 38 | 38 | 73 (16.2) |
|  | Witnessing within the family, relatives, or neighbors | 14 | 14 | 28 (6.2) |
|  | Others (including hospital education, religious houses, and sexual relationships) | 2 | 5 | 7 (1.5) |
| FGM lacks medical reasoning |  | 213 | 206 | 419 (92.7) |
| FGM is still occurring in the community |  | 132 | 98 | 230 (50.9) |
| FGM violates girls' and women's human rights |  | 226 | 215 | 441 (97.6) |
| Forms of violations associated with FGM (Multiple responses allowed) | Intimate partner violence | 138 | 126 | 264 (58.4) |
|  | Child, early, and forced marriage | 138 | 141 | 279 (61.7) |
|  | Stigma and gender discrimination | 136 | 115 | 251 (55.3) |
|  | Right to health, security, and physical integrity | 131 | 118 | 249 (55.1) |
|  | Right to life in cases of death | 144 | 132 | 276 (61.1) |
|  | Right to be free from torture and cruel, inhuman, or other degrading treatment | 73 | 80 | 153 (33.8%) |
| National and international laws criminalize FGM |  | 220 | 217 | 437 (96.7) |
| Aware of 'The International Day of Zero Tolerance for FGM, February 6', and its aim |  | 218 | 206 | 424 (93.8) |
| Desired FGM abandonment |  | 219 | 214 | 433 (95.8) |

### Qualitative findings: lived experiences, systemic barriers, and participant-driven solutions

The ten interviews recruited two participants from each of the five hospitals. Tribes: Seven were Gogo, with Maasai, Burunge, and Rangi (1 each). Ages: 15-16 (2), 17-18 (3 each), and 19 (2). Half were married or unmarried. Six had secondary education; four had primary education. The three main themes are: the hidden system, blood and lies, and intergenerational revolt, supported by six categories and 27 codes (Table 6).

#### Theme 1: The hidden system

FGM persists as a covert, adaptive system where public ceremonies have been abandoned in favor of secretive practices on infants and very young children to evade laws, making it difficult to determine the current rates. As participants described, “now babies are still cut discreetly very young, even before puerperium, is complete” (KI6); “knowing how much, currently, is hard…ceremonies are no longer held…” (KI8); as even “practitioners disguise themselves as traditional healers or birth attendants… their identities are kept classified among a few elders…” (KI10).

Specific gender dynamics reinforce this concealment. While elder women lead the practice, fathers sustain it through passive complicity, often dismissing FGM as a “woman’s issue” (KI3) and upholding the belief it is necessary for marriageability, as they “dare not punish their wives or mothers” (KI9). This highlights a critical need for targeted interventions to build “fathers’ self-efficacy and legitimize their decision-making power over elders’ intentions” (KI8).

To maintain this hidden system, parents actively deceive children and avoid direct conversations, fearing that “if they find out early, they could expose it, cause trouble, or refuse” (KI1). Most (7/10) underwent FGM in early childhood and had no recall of the event. The remaining three, cut at ages 6-8, retained conscious memories. Notably, only three of the ten believed FGM made them “complete women.” Deliberate parental actions ensured the maximum delay in noticing physical differences among girls by forbidding direct naked interaction with other uncut girls; the mother frequently claimed, “Your bodies are clean, keep it secret, meant only for husbands” (KI9). Despite this, seven participants eventually noticed bodily abnormalities; for some, this realization was delayed until adolescence (age 10 or older), prompted by sexual activity or peer exposure, as “at home, all the girls looked the same” (KI6)

Strategies to hide a sibling’s cutting included sending other children away, then returning after it is done to create a fresh environment, with one witness recalling, “They removed us. When we returned, they barred us from seeing her naked. All care was strictly in the room; only adults could care for her. Mother said she was sick, and it was the doctor’s order to prevent contact until she recovered. After three months, we were allowed to discover the healed scar” (KI4). In an unfavorable home environment, a pretence of medicalization was used to take the child away for cutting, claiming “she is sick and going to the traditional healer” (KI5).

#### Theme 2: Blood and lies

For those cut at an age of awareness, the experience was an ‘inhumane process’ of ‘violence disguised as care,’ characterized by force, restraint, and profound pain. The central deception framed this violence as a rite to become ‘complete women.’ Yet these traumatic memories fueled a powerful desire to break the intergenerational cycle, despite powerlessness. A KI recounted her forceful cutting experience:

When I was around six, my mother took me to my grandmother’s village and found two aunties there as well. I didn’t know their plan. The next morning, they woke me up to bathe. A while ago, an unknown older woman came; they boiled medicine and whispered inside for minutes, then I was called. Upon entry, the grandmother said, ‘We are making you a clean and complete woman; you will be cut, do not worry.’ I was scared and tried to run, but I was grabbed. They covered my face and mouth and held me down. The older woman spat the medicine all over my body and forcibly cut me. Nobody came to my aid. I fainted from the pain. When I woke up, they caressed me, saying, ‘You are now a complete woman, the pain will be short, be strong.’ My perineum was full of scars from the forceful cutting. I was in indescribable pain for over a month, especially during urination. The pain I went through was not of this world. I hate everyone every time I think about it… The frequent complaint of my husband’s sexual dissatisfaction and bullying is also killing me. I feel like I came to this world to suffer. At times, I feel miserable and detest my body…the cup I should drink for the rest of my life…Although I lack power…I would not want my child to go through the same cup (moment of sorrow and tears) (KI2).

#### Theme 3: Intergenerational revolt

A central contradiction emerged between participants’ awareness of FGM’s harms and their profound sense of powerlessness to stop it. This “knowledge-power gap” stems from a lack of authority, pressure to uphold norms, lack of uncertainty, economic dependence, and fear of social sanctions. As one explained, “I cannot stop the elders, if it’s to follow traditions…I don’t even remember what was done to me” (KI2), while another noted that young husbands, seen as “children with nothing,” also lack the power to object (KI3).

Participants identified that effective abandonment requires moving beyond individual awareness to address these systemic barriers. Their recommendations form a multi-pronged strategy: (1) Prioritize early, youth-focused education, as “children’s perspectives are more malleable” (KI7); (2) Transform family and community engagement by ensuring “education combines partner involvement and extends to age-based community conferences” to build a united front (KI10, 17); (3) Alternative rites by marrying later with financial independence to “gain strength” (KI3); and Enforce legal accountability and proactive surveillance. They argued for a “strategy focused on law enforcement and investigating all children” (KI3) and proposed “a tracking policy of all children from now on, with the option to sue offenders even after 20 years” to establish discouragement and accurately determine current prevalence (KI8).

Lastly, they highlighted the need to localize global efforts, with one noting she had “heard about World FGM Day but never knew what it exactly meant” (KI1), highlighting the gap between international campaigns and community-level awareness.

## Discussion

This mixed-methods quasi-experimental study demonstrates that a theory-informed, community-based intervention significantly improved FGM awareness and attitudes among young adults in a high-prevalence Tanzanian district. The study tested the causal pathway proposed by our integrated behavioral theories. Implemented with high fidelity, it achieved strong engagement and reach across schools, hospitals, and communities, linking activities to outcomes. A key indicator of this engagement’s sustainability is that the champion network, organized through a WhatsApp group and health clubs, remained active for more than a year post-intervention, transitioning into a community-led platform for ongoing FGM abandonment activities.

The significant quantitative changes confirm the intervention’s success in affecting individual-level mediators (knowledge, perceived risk, and intention). However, the synthesis of quantitative, qualitative, and prevalence data revealed a complex reality: a resilient “hidden system” of practice and a critical “knowledge-power gap.” The qualitative findings demonstrate that these individual-level changes are insufficient to overcome the community-level structural barriers (“the hidden system”), indicating a breakdown in the pathway from individual intention to collective abandonment. This suggests that awareness-raising alone is insufficient without strategies that target structural power dynamics.

### Triangulating evidence: awareness gains amidst a hidden system

The quantitative results are promising: over fivefold increase in adequate awareness (aPR = 5.45) and a near-universal desire for abandonment (95.8%) at endline, aligning with community-engaged strategies [4,22,31]. The integrated theoretical framework likely boosted knowledge, self-efficacy, and social norms. Yet a contradiction of a persistent, adaptive practice remains with a high FGM prevalence (16.6%, 31% among young mothers) and the theme “The Hidden System” explaining the gap with low self-reporting (7.3%). As participants described, practices have shifted to early childhood and deepened into secrecy to evade laws [32]. This covert adaptation is quantified by over 60% of youth aware of FGM’s continuation, who could not explain why it persists, reflecting the powerful social norms that silence discussion and normalize practices [33]. This suggests that recent laws and awareness campaigns have failed to protect this generation, highlighting the need for targeted, youth-engaged interventions such as this study.

### The knowledge-power gap: when awareness lacks agency

The critical synthesis emerges from the tension between improved attitudes and the ‘Intergenerational Revolt’ theme. While 95.8% desired abandonment, they described profound powerlessness due to economic dependence, fear of ostracization, and patriarchal control. This “knowledge-power gap” indicates that the primary barrier is not intention but a lack of executable agency [34]. Mixed-methods insights revealed that familiarity with FGM continuation and knowledge of harms are weakly linked (φ = .114), suggesting awareness alone in a normalized “hidden system” does not prompt clinical enquiry into associated consequences, a cognitive barrier our intervention aimed to address. Additionally, men were 2.35 times more likely to acknowledge FGM’s continuation than women. Qualitative data explained this through patriarchal delegation: fathers are passively complicit, viewing FGM as “a woman’s role,” allowing men to acknowledge it more openly due to perceived low risk. In contrast, women face social pressure to conceal it.

### Implications for policy and practice: a multi-level abandonment strategy

Our integrated findings argue for an evolution in abandonment strategies that moves beyond isolated awareness campaigns:

1. Bridge the gap with empowered participation: programs should combine education with activities that foster youth agency, like economic skill, legal literacy, and leadership. Engaging men and boys as accountable stakeholders, not just allies, is vital to dismantling patriarchal norms that uphold the practice [35,36].
2. Adapt interventions to a covert reality: as FGM becomes more medicalized and secretive [32], protection mechanisms must evolve. This includes training health workers to identify and counsel at-risk families and to create safe, confidential reporting channels that protect whistleblowers.
3. Implement a synchronized ecological approach: sustainable abandonment requires coordinated actions across all levels, child surveillance and punishment (national), legal enforcement and local accountability (societal/community), intergenerational dialogue (interpersonal), and integrating FGM care and education into schools and health services (individual) [16].

### Strengths, limitations, and generalizability

Key strengths include the mixed-methods design, community-based participatory approach, theory-informed intervention, and intervention-naïve sample. The main limitation is the single-arm, baseline-endline design, which cannot definitively establish causality; observed changes may be influenced by secular trends, social desirability bias, or community-wide diffusion of educational messages. Because this was a community-wide campaign with multiple exposure points, we did not track individual intervention attendance, precluding a dose-response analysis. Future efficacy studies could implement controlled exposure tracking. The qualitative findings, though in-depth, are solely from FGM-positive young mothers and lack perspectives of other key groups (e.g., uncut women, elders, practitioners, men).

Our findings’ generalizability is contextual. The sample, selected through a rigorous multi-stage strategy in purposively chosen wards, reflects key demographics of this high-prevalence district. The intervention model, which uses public institutions, is suitable for similar Tanzanian settings, with its feasibility supported by a high retention rate (96.6%).

However, because wards were purposively selected, caution is warranted when applying exact effect sizes more broadly. Nonetheless, the core finding, that awareness can confront resilient structural barriers, likely applies to other contexts in which FGM is a hidden norm, subject to tailored strategies.

## Conclusion

This study showed that a youth-focused, theory-based program can change knowledge and attitudes where none previously existed. It also uncovered the practice’s hidden adaptive nature and how structural powerlessness can weaken positive attitudes. To abandon FGM, a dual approach is needed: ongoing education and empowerment of youth, along with dismantling covert structures through legal, economic, and community actions.

## Abbreviations

aOR: Adjusted Odds Ratio
aPR: Adjusted Prevalence Ratio
CI: Confidence Interval
FGM: Female Genital Mutilation
KI: Key Informant
WHO: World Health Organization.

## Declarations

### Ethical considerations

This study was approved by the College Research Ethics Review Committee of Kilimanjaro Christian Medical University College (Cert. No. 2625). Permissions were obtained from local authorities and institutions. A tiered consent model was used: parental permission and student assent were obtained for school-based activities involving minors under 18; adults provided written consent. For the FGM prevalence audit, trained midwives obtained documented oral consent from mothers to ensure confidentiality and practicality in the high-volume clinical setting. For young mothers (hospital survey/interviews), written consent was obtained; a waiver was granted for mature minors (ages 15-17) to protect them from forced disclosure and to account for the impracticality of contacting parents during short postpartum stays. Household visits required written permission from the head and verbal consent from members. Attendance at public gatherings and hospital training constituted consent. Participation was voluntary, with the right to withdraw. Psychological support was available, and no adverse events occurred. Confidentiality was maintained through the use of anonymized codes, secure storage, and deletion of identifying links after analysis.

### Consent for publication

Not applicable

### Author contributions

LBK and MNI: Investigation, Data Curation, Formal Analysis, and Writing – Original Draft. BZT and ACM: Validation of Analysis and Writing – Review & Editing. BTM: Supervision, Funding Acquisition, and Critical Appraisal of Draft. However, all authors conceptualized and approved the final draft.

## Data Availability

Due to the sensitive nature of this research and to protect participant confidentiality as specified in the ethical approvals, the underlying datasets are not publicly available. This includes the raw qualitative interview transcripts and the primary hospital audit data. The quantitative survey dataset is available from the corresponding author upon reasonable request, subject to a data use agreement and approval from the Kilimanjaro Clinical Research Institute (KCRI). The minimal dataset necessary to replicate all key findings, including the aggregated prevalence data presented in Table 5, is provided within this article and its supplementary materials.

## Acknowledgements

We thank all members of the Chamwino community for their personal support and involvement in the study.

## Data availability statement

**Table 5.** Observed FGM prevalence, types, and tribal distribution among delivering mothers in Chamwino District hospitals, June 2023 – February 2024.

| Hospital |  |  |  |  |  |  |
| --- | --- | --- | --- | --- | --- | --- |
| Metric | Chamwino District Hospital | Dabalo Health Centre | Haneti Health Centre | Mpwayungu Health Centre | Chamwino Health Centre | Total |
| All mothers |  |  |  |  |  |  |
| Total mothers (N) | 1168 | 248 | 524 | 568 | 1262 | 3770 |
| Total FGM, n (%) | 132 (11.3) | 40 (16.1) | 270 (51.5) | 122 (21.5) | 60 (4.8) | 624 (16.6) |
| Young mothers (15-19 years) |  |  |  |  |  |  |
| Total Mothers (N) | 161 | 18 | 75 | 76 | 180 | 510 |
| FGM cases, n (%) | 42 (26.1) | 6 (33.3) | 56 (74.7) | 18 (23.7) | 36 (20.0) | 158 (31.0) |
| FGM typology (All positive mothers) |  |  |  |  |  |  |
| Type I, n | 12 | - | 4 | - | - | 16 |
| Type II, n (%) | 120 (90.9) | 40 (100) | 40 (100) | 122 (100) | 60 (100) | 608 (97.4) |
| Tribal distribution (All FGM cases) |  |  |  |  |  |  |
| Gogo, n (%) |  |  |  |  |  | 459 (73.6) |
| Others, n (%) |  |  |  |  |  | 165 (26.4) |

**Table 6.**
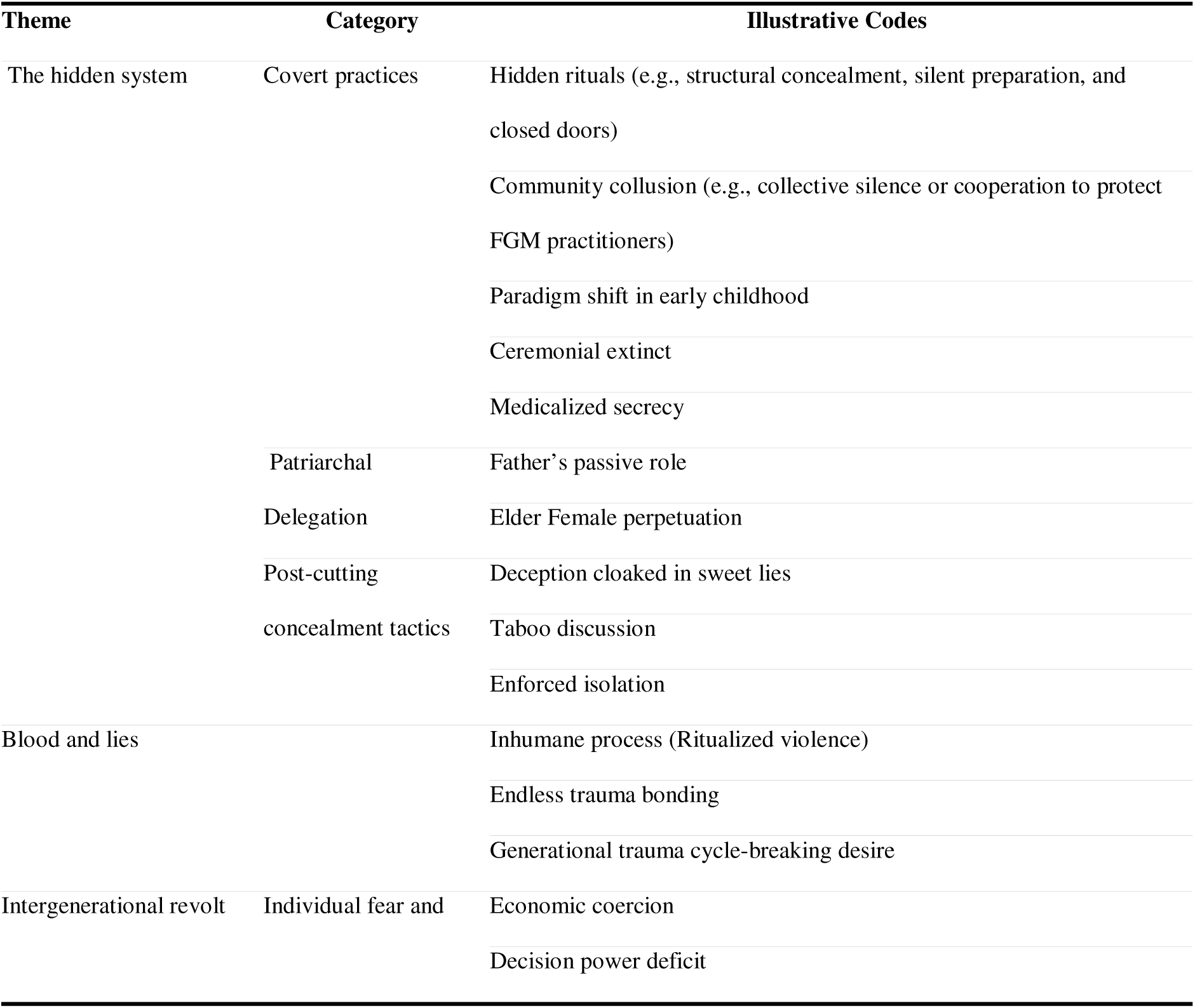
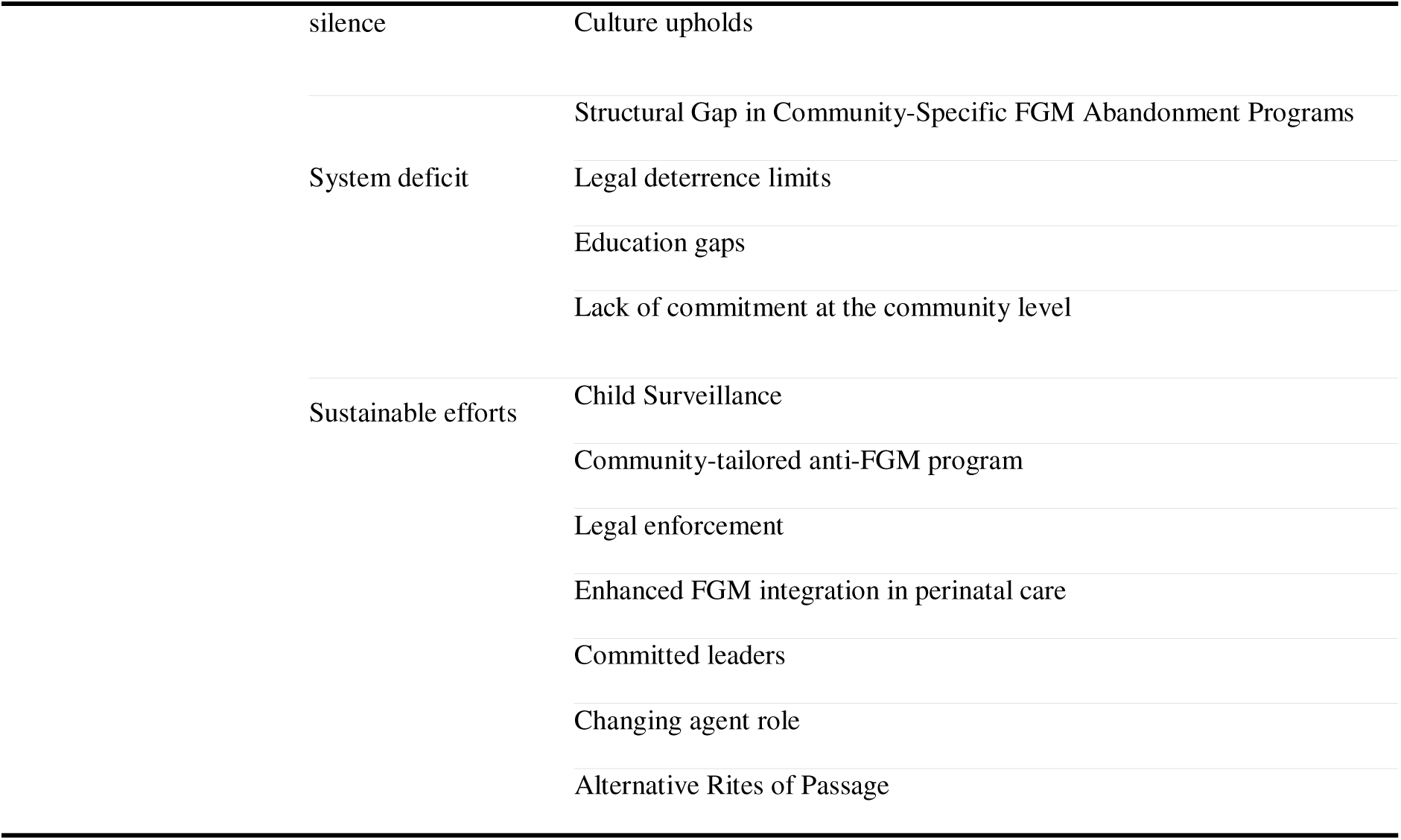
Themes, Categories, and Codes.

## Supporting information

S1 Table. Socio-demographic characteristics of all young adult participants at baseline (N=468), Chamwino District, Tanzania.

S1 File. TREND Statement Checklist

S2 File. COREQ Checklist

S3 File. Data Collection Instruments

S4 File. Training Manual

