## Supplementary material for "Targeting a future generation free from female genital mutilation: a mixed-methods quasi-experimental study of an awareness intervention in central Tanzania": S1_FILE.pdf

### S1 File: TREND Statement Checklist

| Paper Section/ Topic | Item No | Descriptor | Reported? ✓ | Pg # |
|---|---|---|---|---|
| **Title and Abstract** | | | | |
| Title and Abstract | 1 | • Information on how unit were allocated to interventions | ✓ | Title page, 1 |
| | | • Structured abstract recommended | ✓ | Abstract, 2 |
| | | • Information on target population or study sample | ✓ | Abstract, 2 |
| **Introduction** | | | | |
| Background | 2 | • Scientific background and explanation of rationale | ✓ | Introduction (3-4) |
| | | • Theories used in designing behavioral interventions | ✓ | Theoretical framework, 5 |
| **Methods** | | | | |
| Participants | 3 | • Eligibility criteria for participants, including criteria at different levels in recruitment/sampling plan (e.g., cities, clinics, subjects) | ✓ | Participants' sampling, recruitment, and eligibility criteria (6-8) |
| | | • Method of recruitment (e.g., referral, self-selection), including the sampling method if a systematic sampling plan was implemented | ✓ | Participants' sampling, recruitment, and eligibility criteria (6-8) |
| | | • Recruitment setting | ✓ | Study setting and participants (5-8) |
| | | • Settings and locations where the data were collected | ✓ | Study setting and participants (5-8) |
| Interventions | 4 | • Details of the interventions intended for each study condition and how and when they were actually administered, specifically including: | ✓ | Interventions (8-9) |
| | | ○ Content: what was given? | ✓ | Interventions (8-9) |
| | | ○ Delivery method: how was the content given? | ✓ | Interventions (8-9) |
| | | ○ Unit of delivery: how were the subjects grouped during delivery? | ✓ | Interventions (8-9) |
| | | ○ Deliverer: who delivered the intervention? | ✓ | Interventions (8-9) |
| | | ○ Setting: where was the intervention delivered? | ✓ | Interventions (8-9) |
| | | ○ Exposure quantity and duration: how many sessions or episodes or events were intended to be delivered? How long were they intended to last? | ✓ | Interventions (8-9) |
| | | ○ Time span: how long was it intended to take to deliver the intervention to each unit? | ✓ | Interventions (8-9) |
| | | ○ Activities to increase compliance or adherence (e.g., incentives) | ✓ | Interventions (8-9) |
| Objectives | 5 | • Specific objectives and hypotheses | | Study design and registration, 4 |
| Outcomes | 6 | • Clearly defined primary and secondary outcome measures | ✓ | Outcomes and measurement (9-10) |
| | | • Methods used to collect data and any methods used to enhance the quality of measurements | ✓ | Data collection procedures (10-11) |
| | | • Information on validated instruments such as psychometric and biometric properties | ✓ | Data collection procedures (10-11) |
| Sample Size | 7 | • How sample size was determined and, when applicable, explanation of any interim analyses and stopping rules | ✓ | Participants' sampling, recruitment, and eligibility criteria (6-8) |

**S1 File: TREND Statement Checklist**

| | | | | |
|---|---|---|---|---|
| Assignment Method | 8 | • Unit of assignment (the unit being assigned to study condition, e.g., individual, group, community) | ✓ | Participants' sampling, recruitment, and eligibility criteria (6-8) |
| | | • Method used to assign units to study conditions, including details of any restriction (e.g., blocking, stratification, minimization) | ✓ | Participants' sampling, recruitment, and eligibility criteria (6-8) |
| | | • Inclusion of aspects employed to help minimize potential bias induced due to non-randomization (e.g., matching) | ✓ | Participants' sampling, recruitment, and eligibility criteria (6-8) |
| Blinding (masking) | 9 | • Whether or not participants, those administering the interventions, and those assessing the outcomes were blinded to study condition assignment; if so, statement regarding how the blinding was accomplished and how it was assessed. | N/A | N/A |
| Unit of Analysis | 10 | • Description of the smallest unit that is being analyzed to assess intervention effects (e.g., individual, group, or community) | ✓ | Data analysis (11-13) |
| | | • If the unit of analysis differs from the unit of assignment, the analytical method used to account for this (e.g., adjusting the standard error estimates by the design effect or using multilevel analysis) | ✓ | Data analysis (11-13) |
| Statistical Methods | 11 | • Statistical methods used to compare study groups for primary methods outcome(s), including complex methods of correlated data | ✓ | Data analysis (11-13) |
| | | • Statistical methods used for additional analyses, such as a subgroup analyses and adjusted analysis | ✓ | Data analysis (11-13) |
| | | • Methods for imputing missing data, if used | ✓ | Data analysis (11-13) |
| | | • Statistical software or programs used | ✓ | Data analysis (11-13) |
| **Results** | | | | |
| Participant flow | 12 | • Flow of participants through each stage of the study: enrollment, assignment, allocation, and intervention exposure, follow-up, analysis (a diagram is strongly recommended) | ✓ | 8 (Fig 1) |
| | | ○ Enrollment: the numbers of participants screened for eligibility, found to be eligible or not eligible, declined to be enrolled, and enrolled in the study | ✓ | 8 (Fig 1) |
| | | ○ Assignment: the numbers of participants assigned to a study condition | ✓ | 8 (Fig 1) |
| | | ○ Allocation and intervention exposure: the number of participants assigned to each study condition and the number of participants who received each intervention | ✓ | 8 (Fig 1) |
| | | ○ Follow-up: the number of participants who completed the follow- up or did not complete the follow-up (i.e., lost to follow-up), by study condition | ✓ | 8 (Fig 1) |
| | | ○ Analysis: the number of participants included in or excluded from the main analysis, by study condition | ✓ | 8 (Fig 1) |

### S1 File: TREND Statement Checklist

| | | | | |
|---|---|---|---|---|
| | | • Description of protocol deviations from study as planned, along with reasons | ✓ | 8 (Fig 1) |
| Recruitment | 13 | • Dates defining the periods of recruitment and follow-up | ✓ | Participants' sampling, recruitment, and eligibility criteria, 6 |
| Baseline Data | 14 | • Baseline demographic and clinical characteristics of participants in each study condition | ✓ | S1 Table |
| | | • Baseline characteristics for each study condition relevant to specific disease prevention research | ✓ | S1 Table |
| | | • Baseline comparisons of those lost to follow-up and those retained, overall and by study condition | ✓ | Attrition analysis, 15 |
| | | • Comparison between study population at baseline and target population of interest | ✓ | Discussion, 26 |
| Baseline equivalence | 15 | • Data on study group equivalence at baseline and statistical methods used to control for baseline differences | N/A) | |
| Numbers analyzed | 16 | • Number of participants (denominator) included in each analysis for each study condition, particularly when the denominators change for different outcomes; statement of the results in absolute numbers when feasible | ✓ | Fig 1 & Results (13, 19,20) |
| | | • Indication of whether the analysis strategy was "intention to treat" or, if not, description of how non-compliers were treated in the analyses | ✓ | 11-12 (Data analysis) |
| Outcomes and estimation | 17 | • For each primary and secondary outcome, a summary of results for each estimation study condition, and the estimated effect size and a confidence interval to indicate the precision | ✓ | Results (15-17) |
| | | • Inclusion of null and negative findings | ✓ | Results (15, 16) |
| | | • Inclusion of results from testing pre-specified causal pathways through which the intervention was intended to operate, if any | ✓ | Results (13-23) Discussion 24 |
| Ancillary analyses | 18 | • Summary of other analyses performed, including subgroup or restricted analyses, indicating which are pre-specified or exploratory | ✓ | Data analysis (11-13) Results, 15 |
| Adverse events | 19 | • Summary of all important adverse events or unintended effects in each study condition (including summary measures, effect size estimates, and confidence intervals) | ✓ | Ethical considerations, 28 |
| **DISCUSSION** | | | | |
| Interpretation | 20 | • Interpretation of the results, taking into account study hypotheses, sources of potential bias, imprecision of measures, multiplicative analyses, and other limitations or weaknesses of the study | ✓ | 24-27 |
| | | • Discussion of results taking into account the mechanism by which the intervention was intended to work (causal pathways) or alternative mechanisms or explanations | ✓ | 24 |
| | | • Discussion of the success of and barriers to implementing the intervention, fidelity of implementation | ✓ | 24 |
| | | • Discussion of research, programmatic, or policy implications | ✓ | 26 |

### S1 File: TREND Statement Checklist

| | | | | |
|---|---|---|---|---|
| Generalizability | 21 | • Generalizability (external validity) of the trial findings, taking into account the study population, the characteristics of the intervention, length of follow-up, incentives, compliance rates, specific sites/settings involved in the study, and other contextual issues | ✓ | 26-27 |
| Overall Evidence | 22 | • General interpretation of the results in the context of current evidence and current theory | ✓ | 26-27 |

*From:* Des Jarlais, D. C., Lyles, C., Crepaz, N., & the Trend Group (2004). Improving the reporting quality of nonrandomized evaluations of behavioral and public health interventions: The TREND statement. *American Journal of Public Health*, 94, 361-366. For more information, visit: http://www.cdc.gov/trendstatement/
