## Supplementary material for "Targeting a future generation free from female genital mutilation: a mixed-methods quasi-experimental study of an awareness intervention in central Tanzania": S1_Table.pdf

**S1 Table. Socio-demographic characteristics of all young adult participants at baseline (N=468), Chamwino District, Tanzania.**

**Manuscript: Targeting a future generation free from female genital mutilation: a mixed-methods quasi-experimental study of an awareness intervention in central Tanzania**

| Variable (N=468) | | (n, %) | Subtotal (n, %) | Median (Range) |
|---|---|---|---|---|
| **Age (years)** | | | | |
| 15 | | 89 (19.0) | | |
| 16 | | 107 (22.9) | | |
| 17 | | 112 (23.9) | | **17 (15-19)** |
| 18 | | 107 (22.9) | | |
| 19 | | 53 (11.3) | | |
| **Years of community residence** | | | | **16 (1-19)** |
| **Sex** | | | | |
| Male | | 234 (50) | | |
| Female | | 234 (50) | | |
| **Ward of residence** | | | | |
| Mpwayungu | | 8 (1.7) | | |
| Haneti | | 116 (24.8) | | |
| Chamwino | | 116 (24.8) | | |
| Itiso | | 106 (22.6) | | |
| Mlowa | | 8 (1.7) | | |
| Dabalo | | 114 (24.4) | | |
| **Ward type** | | | | |
| Semi-urban | | 124 (26.5) | | |
| Remote | | 344 (73.5) | | |
| **Setting** | | | | |
| | Itiso | 106 (22.6) | | |

| | | | |
|---|---|---|---|
| Secondary school (students) | Chamwino | 107 (22.9) | 426 (91.0) |
| | Haneti | 107 (22.9) | |
| | Dabalo | 106 (22.6) | |
| Hospitals (young mothers) | Mpwayungu Health Centre | 9 (1.9) | 42 (9.0) |
| | Chamwino District Hospital | 8 (1.7) | |
| | Chamwino Health Centre | 9 (1.9) | |
| | Haneti Health Centre | 8 (1.7) | |
| | Dabalo Health Centre | 8 (1.7) | |
| **Education level** | | | |
| Secondary | | 433 (92.5) | |
| Primary | | 35 (7.5) | |
| **Tribe (n =33)** | | 267 (57.1) | |
| Gogo | | 201 (42.9) | |
| Others | | | |
| **Involvement in the anti-FGM program** | | No | 468 (100%) |
