## Supplementary material for "Targeting a future generation free from female genital mutilation: a mixed-methods quasi-experimental study of an awareness intervention in central Tanzania": S2_File .pdf

**S2 File: Consolidated criteria for reporting qualitative studies (COREQ): 32-item checklist**

| **No. Item** | **Guide questions/description** | **Reported on Page #** |
|---|---|---|
| **Domain 1: Research team and reflexivity** | | |
| *Personal Characteristics* | | |
| 1. Interviewer/facilitator | Which author/s conducted the interview or focus group? | 10-11 (Data collection procedures) |
| 2. Credentials | What were the researcher's credentials? E.g., PhD, MD | 1 (Authors' affiliation & Information) |
| 3. Occupation | What was their occupation at the time of the study? | 1 (Authors' affiliation & Information) |
| 4. Gender | Was the researcher male or female? | 1 (Authors' affiliation & Information) |
| 5. Experience and training | What experience or training did the researcher have? | 1 (Authors' affiliation & Information) |
| *Relationship with participants* | | |
| 6. Relationship established | Was a relationship established prior to study commencement? | 4 (Study design and registration)<br>8-9 (Interventions)<br>28 (Ethical Considerations) |
| 7. Participant's knowledge of the interviewer | What did the participants know about the researcher? e.g., personal goals, reasons for doing the research | 8-9 (Interventions)<br>28 (Ethical Considerations) |
| 8. Interviewer characteristics | What characteristics were reported about the interviewer/facilitator? e.g., Bias, assumptions, reasons, and interests in the research topic | 10-11 (Data collection procedures) |

| | | |
|---|---|---|
| **Domain 2: study design** | | |
| *Theoretical framework* | | |
| 9. Methodological orientation and Theory | What methodological orientation was stated to underpin the study? e.g. grounded theory, discourse | 4 (Study design and registration)<br>7 (Participants' sampling, recruitment, and eligibility criteria) |

| | | |
|---|---|---|
| | analysis, ethnography, phenomenology, content analysis | |
| *Participant selection* | | |
| 10. Sampling | How were participants selected? e.g. purposive, convenience, consecutive, snowball | 6-8 (Participants' sampling, recruitment, and eligibility criteria) |
| 11. Method of approach | How were participants approached? e.g. face-to-face, telephone, mail, email | 11 (Data collection procedures) |
| 12. Sample size | How many participants were in the study? | Fig 1<br>7 (Participants' sampling, recruitment, and eligibility criteria) |
| 13. Non-participation | How many people refused to participate or dropped out? Reasons? | 7 (Participants' sampling, recruitment, and eligibility criteria) |
| *Setting* | | |
| 14. Setting of data collection | Where was the data collected? e.g., home, clinic, workplace | 5-6 (Study setting and participants) |
| 15. Presence of non-participants | Was anyone else present besides the participants and researchers? | 11 (Data collection procedures) |
| 16. Description of sample | What are the important characteristics of the sample? e.g., demographic data, date | 20 (Qualitative Findings: Lived Experiences, Systemic Barriers, and Participant-Driven Solutions) |
| *Data collection* | | |
| 17. Interview guide | Were questions, prompts, guides provided by the authors? Was it pilot tested? | 11 (Data collection procedures) |

| | | |
|---|---|---|
| 18. Repeat interviews | Were repeat interviews carried out? If yes, how many? | 11 (Data collection procedures) |
| 19. Audio/visual recording | Did the research use audio or visual recording to collect the data? | 11 (Data collection procedures) |
| 20. Field notes | Were field notes made during and/or after the interview or focus group? | 11 (Data collection procedures) |
| 21. Duration | What was the duration of the interviews or focus group? | 11 (Data collection procedures) |
| 22. Data saturation | Was data saturation discussed? | 7-8 (Participants' sampling, recruitment, and eligibility criteria) |
| 23. Transcripts returned | Were transcripts returned to participants for comment and/or correction? | 11 (Data collection procedures) |
| **Domain 3: analysis and findings** | | |
| *Data analysis* | | |
| 24. Number of data coders | How many data coders coded the data? | 12 (Data analysis) |
| 25. Description of the coding tree | Did authors provide a description of the coding tree? | 12 (Data analysis) |
| 26. Derivation of themes | Were themes identified in advance or derived from the data? | 12 (Data analysis) |
| 27. Software | What software, if applicable, was used to manage the data? | 12 (Data analysis) |
| 28. Participant checking | Did participants provide feedback on the findings? | NA |
| *Reporting* | | |
| 29. Quotations presented | Were participant quotations | 21-24 (Qualitative Findings: Lived |

| | | |
|---|---|---|
| | presented to illustrate the themes/findings? Was each quotation identified? e.g. participant number | Experiences, Systemic Barriers, and Participant-Driven Solutions) |
| 30. Data and findings consistent | Was there consistency between the data presented and the findings? | 20-24 (Qualitative Findings: Lived Experiences, Systemic Barriers, and Participant-Driven Solutions) |
| 31. Clarity of major themes | Were major themes clearly presented in the findings? | 20-24 (Qualitative Findings: Lived Experiences, Systemic Barriers, and Participant-Driven Solutions) |
| 32. Clarity of minor themes | Is there a description of diverse cases or discussion of minor themes? | 20-24 (Qualitative Findings: Lived Experiences, Systemic Barriers, and Participant-Driven Solutions) |
