## Supplementary material for "Targeting a future generation free from female genital mutilation: a mixed-methods quasi-experimental study of an awareness intervention in central Tanzania": S3_File.pdf

**S3 File: Data Collection Instruments**

| | | | |
|---|---|---|---|
| **INSTRUMENT 1:** | **Knowledge, Attitudes, and Perceptions on Female Genital Mutilation (FGM) Structured Questionnaire (English Version)** | | |
| **Participant Code Number** | **Fill your assigned number from (P1 to P468)** | | |
| **Instructions** | **Tick (v) or fill where applicable** | | |
| **PART A: Participants' Demographics** | | | |
| **SN** | **PARTICULAR** | | **DESCRIPTION (If any)** |
| 1 | Your age (in years) | | |
| 2 | Sex | | **[ ]** Male **[ ]** Female |
| 3 | Your ward of residence (mention name) | | |
| 4 | Type of ward | | [ ] Semi-urban [ ] Remote |
| 5 | Setting | School | Name: |
| | | Hospital | Name: |
| 6 | Years of community residence | | |
| 7 | Education level | | [ ] College/University<br>[ ] Secondary<br>[ ] Primary<br>[ ] Not attended |
| 8 | Your tribe | | |
| **PART B: Awareness: Knowledge, Attitudes, and Perceptions on Female Genital Mutilation** | | | |
| 1 | FGM is the partial or total removal of external female genitalia and infliction of other injuries to female genitalia for non-medical reasons | | [ ] Yes [ ] No |
| 2 | Source of information and awareness regarding FGM?<br>(tick all that apply)* | | [ ] Community discussions*<br>[ ] School education*<br>[ ] Mass media*<br>[ ] Witness within family, relatives or neighbors*<br>[ ] FGM positive status (women only)*^<br>[ ] Previous involved in any anti-FGM program*^<br>Others (specify)…………………………* |
| 3 | Is FGM harmful? | | **[ ] Yes [ ] No** |
| 4 | If yes in question 3 above, what could be the side effects of FGM?<br>(Tick (v) all that apply) | | [ ] Infections<br>[ ] Excessive bleeding<br>[ ] Severe pain |

| | | |
|---|---|---|
| | | [ ] Difficult childbirth<br>[ ] Urinary problems<br>[ ] Sexual problems<br>[ ] Psychological problems<br>Others (specify)……………………* |
| 5 | Is FGM still occurring in the community?* | [ ] Yes [ ] No |
| 6 | If yes in question 5 above, what could be the associated sustaining reasons?* | |
| 7 | Does FGM violate the human rights of girls and women? | [ ] Yes [ ] No |
| 8 | If yes in question 7 above, what could be the human rights violation associated with FGM?<br>(Tick (v) all that apply) | [ ] Intimate partner violence<br>[ ] Child, early and forced marriage<br>[ ] Stigma and gender discrimination<br>[ ] Right to health, security, and physical integrity<br>[ ] Right to life in cases of death<br>[ ] Right to be free from torture and cruel, inhuman or other degrading treatment<br>Others (specify)……………………. * |
| 9 | Is FGM a criminal offense by Tanzania and international laws? | [ ] Yes [ ] No |
| 10 | Are you aware of the 'International Day of Zero Tolerance for FGM, February 6', and its aim? | [ ] Yes [ ] No |
| 11 | Do you wish for FGM abandonment? | [ ] Yes [ ] No |
| 12 | Do you feel this project belongs to you/your community? (endline)*^ | [ ] Yes [ ] No |

**Key:** *Excluded from awareness level estimation and paired analysis; ^Explored only at baseline (no need for comparison).

**Exclusion Justification:** (1) Sources of information measure exposure pathways and personal history rather than factual knowledge, (2) 'Is FGM still occurring in the community?' assesses perception of a covert practice influenced by social exposure and length of community residency, rather than objective knowledge, (3) Self-reported FGM positive status (for female participants) and prior involvement in anti-FGM programs were non-universal terms, measuring personal history and exposure, not the current state of knowledge which the score aimed to capture, (4) 'Do you feel this project belongs to you/your community?' was a measure of 'Intervention reach, fidelity, and process evaluation', rather than knowledge, and (5) The open-ended 'Others (specify)' response option represents a non-standardized, qualitative data point that could not be meaningfully scored or compared across participants within the quantitative scoring framework. All excluded items were analyzed separately as descriptive or secondary outcomes.

| **INSTRUMENT 2** | **PART B: Chamwino District FGM Prevalence Clinical Audit Checklist (2023-2024)** |
|---|---|

**Instructions for Nurse-Midwife/ Data Collector:**

1. Complete this checklist for **every** mother following delivery and initial postpartum assessment.
2. Obtain **oral consent** (Section 1) before proceeding.
3. Fill in all applicable fields. Use BLOCK LETTERS.
4. File completed checklist securely in the designated study box at the nurse's station.

**Section 1: Statement of Oral Consent and Data Collector Information**

*Read the following statement to the mother:*
*"Hello. As part of a routine health system check to improve services for all women, we are documenting information about mothers' health during childbirth. This includes checking for any past procedures on the female genitalia. Your information will be kept completely confidential and used only to understand community health needs. Your participation is voluntary. Do you agree for me to include your anonymous information in this check?"*

**Mother Agreement:** [ ] YES [ ] NO
*(If "NO," thank the mother and STOP here. Do not complete the rest of the form.)*

**Data Collector's Name:** ____________________________________________

**Signature:** ___________________________ **Date:** //202___

| **Section 2: Patient General Information for Registry Verification**<br>*(To be completed for all consented mothers)* | | |
|---|---|---|
| **Field** | **Instruction** | **Entry** |
| **Facility Code** | *Use your facility's code (see box).* | **[ ] CDH [ ] CHC [ ] MPH [ ] DHC [ ] HHC** |
| **Date of Delivery** | DD/MM/YYYY | __ / __ / 202__ |
| **Shift at Delivery** | Tick one. | **[ ] Morning [ ] Evening [ ] Night** |
| **Maternal Age** | In completed years. | __ years |
| **Sequential Birth Number** | This mother's delivery number for today at this facility (e.g., 1st=1, 2nd=2). | __ |
| **De-identified Mother Code (UID)** | *Create code using formula:* **[Facility Code] - [Last 2 of Year][Month][Day] - [Seq. Birth No.]** *Example: 5th delivery at CHC on Oct 25, 2023 =* **CHC-231025-05** | ________-______-__ |
| **Section 3: FGM Specific Clinical Data**<br>*(To be completed after physical examination)* | | |
| **Field** | **Instruction** | **Entry** |

| | | |
|---|---|---|
| **FGM Status** | Based on visual examination. | **[ ] Positive [ ] Negative** |
| **If Positive, WHO Type** | *Refer to WHO typology poster.* | **[ ] I [ ] II [ ] III [ ] IV** |
| **Mother's Tribe** *As reported by the mother.* ______________________________ | | |

| | |
|---|---|
| **INSTRUMENT 3** | **Interview Guide for FGM-Positive Young Mothers (English Version)** |
| **Participant Code Number (e.g., KI1, KI2--)** | |
| **PART A: Participants' Demographics** | |
| 1 | Setting (hospital name) |
| 2 | Participant tribe |
| 3 | Participant age (years) |
| 4 | Marital status |
| 5 | Education level |
| Qn1 | Could you tell me about your own experience with FGM? You might think about when it happened, what you remember, and how you felt at that time. |
| Qn2 | From your perspective, how is FGM practiced in the Chamwino community today? What, if anything, has changed compared to the past? |
| Qn3 | In your view, what power do young people or children have to say 'no' to FGM, either for themselves or for their future children? What makes it easy or difficult for them? |
| Qn4 | Thinking about your life now, in what ways, if any, does having undergone FGM affect your daily life, your health, or your relationships? Could you describe an example of a specific challenge or difficulty you have faced? |
| Qn5 | Many people in the community say they know FGM is harmful, yet it continues. In your opinion, what are the most important reasons it persists? |
| Qn6 | What do you see as the most important role for young people, like yourself, in helping the community move away from FGM? What would they need to be able to play that role effectively? |
| Qn7 | If the community decided to end FGM, what do you think are the most practical and effective steps it could take? Who would need to be involved? |
