## Supplementary material for "Targeting a future generation free from female genital mutilation: a mixed-methods quasi-experimental study of an awareness intervention in central Tanzania": S4_File.pdf

**S4 File: Training Manual**

### ENGLISH VERSION

**Aim**

To provide a structured, theory-informed educational resource for training community stakeholders in Chamwino District on Female Genital Mutilation (FGM). The manual aims to increase knowledge of FGM health risks, shift attitudes, and build skills for promoting abandonment, using content derived from global evidence and framed within established behavioral theories (Health Belief Model, Social Cognitive Theory, Theory of Planned Behaviour, Socio-Ecological Model) to address individual, interpersonal, and community-level drivers of the practice.

**Definition, Global Burden, and Local Context of FGM**

FGM is a procedure involving the partial or total removal of external female genitalia or other injury to the female genital organs for non-medical reasons, as defined by the World Health Organization (WHO). It is a harmful practice and a violation of the human rights of girls and women. With origins over two millennia, FGM persists in approximately 31 countries, predominantly in Africa, which accounts for over 80% of global cases. In Tanzania, national prevalence among women of reproductive age is 8%, with stark regional disparities. The Dodoma Region has the country's second-highest prevalence (47%), underscoring its status as a critical priority area for targeted efforts to address abandonment.

**Immediate and Long-Term Health Consequences of FGM**

The health consequences of FGM are severe and multifaceted, affecting individuals, partners, and families. Complications are categorized as immediate or long-term. Immediate risks include severe pain, haemorrhage, shock, infection, urinary retention, and injury to adjacent genital tissue, which can be fatal. Long-term sequelae encompass a broad spectrum of gynecological, obstetric, sexual, and psychological harm. These include chronic pain, dysmenorrhea, dyspareunia (painful intercourse), and reduced sexual satisfaction, and a significantly heightened risk of obstetric complications such as obstructed labour, postpartum haemorrhage, need for surgical delivery, and perinatal mortality. Additionally, women living with FGM are at increased risk of psychological disorders, including depression, anxiety, post-traumatic stress disorder (PTSD), and diminished self-esteem, as well as heightened vulnerability to intimate partner violence.

**WHO Classification and Context of FGM Types**

The WHO classifies FGM into four main types (I–IV), defined by the nature and extent of tissue removed or genital injury inflicted. This typology is rooted in the specific social, cultural, and ritual contexts of practicing communities. The severity of associated immediate and long-term health risks is generally correlated with the extent of cutting, with more extensive procedures (e.g., Type III, infibulation) posing greater obstetric, sexual, and psychological harm. In many settings, Type II (excision of the clitoris and labia minora) is the most commonly reported form of the practice.

*[Note: A clinical illustration of FGM types was removed from this public version due to copyright restrictions. The classification is described in Table 1 below.]*

**Table 1: WHO Classification of Female Genital Mutilation**

| WHO Type | Common Terminology | WHO Definition/ Genital Modification |
|---|---|---|
| **Type I** | Clitoridectomy | Partial or total removal of the clitoris (clitoral glans) and/or the prepuce (clitoral hood). |
| **Type II** | Excision | Partial or total removal of the clitoris and the labia minora, with or without excision of the labia majora. |
| **Type III** | Infibulation | Narrowing of the vaginal orifice with the creation of a covering seal by cutting and appositioning the labia minora and/or labia majora, with or without excision of the clitoris. |
| **Type IV** | Other | All other harmful procedures to the female genitalia for non-medical purposes (e.g., pricking, piercing, incising, scraping, cauterization). |

**Socio-Cultural and Structural Drivers of FGM**

The persistence of FGM is sustained by a complex interplay of deeply entrenched socio-cultural norms, gender inequalities, and economic factors. It is perpetuated across generations as a traditional practice, often without critical examination of its harmful consequences. Key drivers include the desire to control female sexuality, encompassing beliefs about preserving virginity, ensuring marital fidelity, and preventing premarital pregnancy, and to fulfill cultural rites of passage into womanhood. Additional sustaining factors are beliefs about hygiene and aesthetic “cleanliness,” the enhancement of a girl’s marriageability and social status, economic incentives for practitioners, community pressure, and veneration of ancestors. Crucially, a widespread lack of awareness regarding the severe and lifelong health risks associated with FGM enables these misconceptions to prevail.

**Practitioners, Legal Framework, and Evolving Challenges in Tanzania**

FGM varies by context, with some countries seeing medicalization by healthcare professionals, while in Tanzania, it's mainly performed by traditional healers and birth attendants. The Sexual Offenses Special Provisions Act (SOSPA) of 1998, amended in 2016, criminalizes FGM with penalties of 5 to 15 years, or life imprisonment if it causes death. It also imposes penalties on those aiding the practice. Enforcement has made FGM more covert, especially on infants and young children. Strategies to end FGM must involve community-led detection, dismantling practitioner networks, and empowering youth to reject, report, and protect themselves from the practice.

**The Multifaceted Benefits of FGM Abandonment**

Abandoning FGM provides profound benefits for individuals, families, and societies by preventing physical and psychological harm, such as trauma, pain, sexual dysfunction, obstetric issues, and mental health disorders. It upholds human rights related to bodily integrity, security, and freedom from torture, promoting gender equality. Ending this practice breaks intergenerational trauma cycles, supports healthier communities, and allows resource reallocation for development. It also aligns nations with global human rights goals, including Sustainable Development Goal (SDG) 5.3 to eliminate FGM by 2030, boosting their commitment to human dignity.

**Multi-Level Strategies for FGM Abandonment**

Efforts to abandon FGM are made globally, nationally, and locally through various strategies. Globally, initiatives such as the International Day of Zero Tolerance for FGM (February 6) and SDG 5.3 provide a framework and focus. Nationally, laws like Tanzania's Sexual Offences Special Provisions Act establish normative and preventive standards. But the most impactful strategies are community-based, addressing socio-cultural roots through educational campaigns on health risks, promoting alternative rites that maintain cultural identity, and empowering community champions to change norms. Support systems such as healthcare, psychosocial services, legal aid, and advocacy are vital. Successful efforts actively involve community members, especially empowering youth, men, and fathers as allies to abandon FGM.
